# Risk of dementia after hospitalization due to traumatic brain injury: a longitudinal, population-based study

**DOI:** 10.1101/2021.06.20.21259106

**Authors:** Rahul Raj, Jaakko Kaprio, Pekka Jousilahti, Miikka Korja, Jari Siironen

**Affiliations:** Department of Neurosurgery, Helsinki University Hospital and University of Helsinki, Helsinki, Finland; Institute for Molecular Medicine Finland, University of Helsinki, Helsinki, Finland; Department of Public Health and Welfare, Finnish Institute for Health and Welfare, Helsinki, Finland

**Keywords:** traumatic brain injury, dementia, neurodegenerative disease, dementia, epidemiology

## Abstract

**Objective:** Traumatic brain injury (TBI) is considered a potential modifiable dementia risk factor. We aimed to determine whether TBI actually increases the risk of dementia when adjusting for other relevant dementia risk factors.

**Methods:** National prospective longitudinal cohort study that included 32,385 participants aged 25–64 during 1992–2012. Of these, 288 suffered from a major TBI (diagnosis of traumatic intracranial hemorrhage and hospital length of stay (LOS) ≥3 days) and 406 from a minor TBI (concussion diagnosis and hospital LOS ≤1 day). Dementia was defined as any first hospital contact with a diagnosis of dementia; first use of an anti-dementia drug; and dementia as an underlying or contributing cause of death. Follow-up until the end of 2017 yielded a total of 1,010 incident dementia cases.

**Results:** After adjusting for age and sex, hospitalization due to major TBI (hazard ratio [HR] 1.51, 95% CI 1.03–2.22), but not minor TBI, increased the risk of dementia. After additional adjustment for educational status, smoking status, alcohol consumption, physical activity, and hypertension, the association between major TBI and dementia weakened (HR 1.30, 95% CI 0.86–1.97). The risk factors most strongly attenuating the association between major TBI and dementia were alcohol consumption and physical activity.

**Conclusion:** There was an association between major TBI and incident dementia. The association was attenuated after adjusting for confounders, especially alcohol consumption and physical activity. Minor TBI was not associated with an increased risk of dementia.

## INTRODUCTION

Annually, over 27 million people suffer from a traumatic brain injury (TBI), and the incidence is increasing [1]. Recently, TBI was added as a new, potentially specific modifiable risk factor of dementia by the 2020 Lancet Commission on dementia prevention, intervention and care [2]. The same review identified 11 other dementia risk factors that may confound the association of TBI with dementia. Of these, low cognitive performance, less education, substance use (heavy alcohol consumption and smoking) and low physical fitness are associated with TBI, dementia and early death, making it essential to adjust for these when assessing the association between TBI and dementia [3–11]. With the exception of one very recent study [12], no earlier studies have adequately controlled for these potential confounders that are associated with both TBI and dementia (Online Supplemental File 1) [13]. Due to the steadily increasing number of people living with dementia, it is imperative to identify real risk factors in order to maximize the effectiveness of preventive measures [2].

Our aim was to assess the independent association between TBI and dementia while carefully adjusting for other relevant dementia risk factors. We hypothesized that, after confounding adjustment, major TBI—but not minor TBI—would be associated with an increased risk of dementia [14,15]. We also set out to identify confounders affecting the relationship between TBI and dementia.

## METHODS

### FINRISK

The National Institute for Health and Welfare approved the study and granted us access to the FINRISK database (THL/155/6.00.00/2019).

The FINRISK surveys have been described in detail previously [16]. Briefly, national FINRISK surveys have been carried out in five-year intervals since 1972, focusing on independent, population-based, random samples from various geographical areas of Finland. The surveys include a self-administered questionnaire, physical measurements and blood samples. For this study, we obtained data from the surveys conducted in 1992, 1997, 2002, 2007 and 2012. We included participants between 25 and 64 years of age [14]. For those who participated several times, we included the first survey they completed.

### Definition of major and minor traumatic brain injury

We defined minor TBI as an International Statistical Classification of Diseases and Related Health Problems 8 or 9 (ICD-8 or ICD-9) diagnosis of 850 and an ICD-10 diagnosis of S06.0, according to the criteria of the US Centers for Disease Control and Prevention (CDC), with the exception of isolated skull fractures [14,17]. We defined major TBI as an ICD-8 or ICD-9 diagnosis of 851–854 or an ICD-10 diagnosis of S06.1–S06.9. To reduce the likelihood that participants with a minor TBI had a major TBI, we only considered participants hospitalized for one day or less. Similarly, to reduce the likelihood that participants with a major TBI only had a minor TBI, we considered participants hospitalized for three days or longer. If a participant had a history of both minor and major TBI, the major TBI diagnosis was considered. We extracted TBI diagnoses between January 1970 and December 2017 (ICD-8 was in use until 1986, ICD-9 was used from 1987–1995 and ICD-10 was used from 1996 onwards) from the Finnish Care Register.

### Definition of dementia

We defined the date of dementia diagnosis as the date when the participant was first prescribed an anti-dementia drug (Anatomical Therapeutic Chemical Classification System N06D*) or as the first date on which the participant was hospitalized for any reason with a primary or secondary diagnosis of dementia. We also identified participants for whom dementia was recorded on their death certificate as an underlying or contributing cause of death.

We obtained data regarding anti-dementia drug prescriptions through the National Social Insurance Institution’s (Kela) drug register. In Finland, persons diagnosed with Alzheimer’s disease (AD) are granted anti-dementia drug reimbursement given major functional impairment caused by the diagnosis. Diagnosis is based upon clinical neurological examination, cognitive testing and, if necessary, brain imaging. The national Current Care Guidelines for dementia recommend that anti-dementia drugs should be started for all with a new diagnosis of AD, unless there is a contraindication for their use [18]. Reimbursement for anti-dementia drugs started in February 1999, and it is not restricted based on the severity of dementia (i.e. there are no lower or upper limits). We collected data on all new anti-dementia drug prescriptions from 1993 until December 2017.

Regarding hospitalization with a diagnosis of dementia, we defined dementia as an ICD-8 or ICD-9 diagnosis of 331, 290 or 4378A or an ICD-10 diagnosis of G30, F00, F01, F02 or F03 [19]. We extracted diagnoses of hospitalized dementia between January 1970 and December 2017 from the Finnish Care Register. The validity of the Finnish Care Register for dementia diagnoses is good [19].

To minimize the possibility of reverse causality, we only considered dementia diagnoses made one year after TBI [14,20].

### Definition of covariates

Covariates (dementia risk factors [2,10]) associated with TBI were obtained through the FINRISK studies [21]. Educational status was defined as low, middle or high. Smoking status was defined as non-smoker, former smoker (stopped smoking over half a year ago) or current smoker, and we divided current smokers based on the number of cigarettes used per day (≤15 cigarettes/day or >15 cigarettes per day). Alcohol consumption was defined as non-drinker, light drinker, moderate to heavy drinker. Leisure time physical inactivity was defined as sedentary, light activity or moderate to intensive activity. Hypertension was defined based on a systolic blood pressure of >140 mmHg or a diagnosis of hypertension prior to participation. Detailed descriptions and definitions of the covariates can be found in the Online Supplemental File 2.

### Statistical analyses

We used Stata (version 15, StataCorp, College Station, TX) to conduct the statistical analyses.

Participants were considered to enter the study at the time of their participation in a FINRISK study, and participant age was the underlying time parameter. We defined the end of follow-up as death, date of dementia diagnosis or December 31, 2017, whichever came first.

To assess the association between TBI and risk of dementia, we used Cox proportional hazard models. We separately compared participants with a history of minor and major TBI to participants with no TBI. We created a partially adjusted model and a fully adjusted model. In the partially adjusted model, we adjusted for sex and year of FINRISK participation. In the fully adjusted model, we also adjusted for educational status, smoking status, alcohol consumption, leisure time physical activity and hypertension [2,10]. In the primary analysis, all participants with TBI were included, regardless of whether the TBI occurred before or after FINRISK participation.

The results are presented as hazard ratios (HRs), subhazard ratios (sHRs) or odds ratios (ORs) with 95% confidence intervals (CIs). According to the Schoenfeld residuals, our models met the proportional assumption criteria. Because of the low number of missing data per variable, we performed complete case analyses.

We followed the STROBE (Strengthening the Reporting of Observational studies in Epidemiology) statement to report our results [22].

### Sensitivity analyses

TBI, especially major TBI, is associated with a high excess risk of death, and it shares risk factors with dementia [3,8,9,23,24]. Thus, participants with a history of TBI prior to FINRISK participation represent a select cohort of participants who survived the initial TBI (and thus were able to participate). Therefore, we performed a sensitivity analysis that included only those who were hospitalized due to a TBI after FINRISK participation. Further, we conducted another sensitivity analysis to evaluate the association between TBI and dementia using a competing risks model [25].

### Nested cohort analyses

We also conducted two nested cohort analyses. In the first (case–control), we matched participants by analysis time (*sttocc* in Stata, 4 controls and 1 case) and sex. We then assessed the association between TBI and dementia using conditional logistic regression, adjusting for educational status, smoking status, alcohol consumption, leisure time physical activity and hypertension. In the second analysis (exposed–nonexposed), we separately matched up to two non-TBI controls with participants with minor TBI and participants with major TBI by sex, educational status, smoking status, alcohol consumption, leisure time physical activity and hypertension (*ccmatch* in Stata). We then assessed the association between TBI and dementia in a matched Cox proportional hazard model, using age as the underlying parameter.

## RESULTS

The study population constituted 32,385 individuals (Online Supplemental Table 1). The total time at risk was 500,954 person-years (median 15.8 years). Of the total population, 288 participants (0.9%) had a history of major TBI (127 participants before FINRISK participation) and 406 (1.3%) had a history of minor TBI (238 participants before FINRISK participation) without a history of dementia at baseline or within one year of injury (Figure 1). There were no major differences in the risk factors for participants with TBI before or after FINRISK participation (Online Supplemental Table 2). The median age at the time of minor TBI was 40.1 years, compared to 54.3 years for major TBI. Females comprised a minority of participants with major TBI (28.5%), but 41.4% and 53.9% of those with minor or no TBI, respectively.

**Figure 1:**
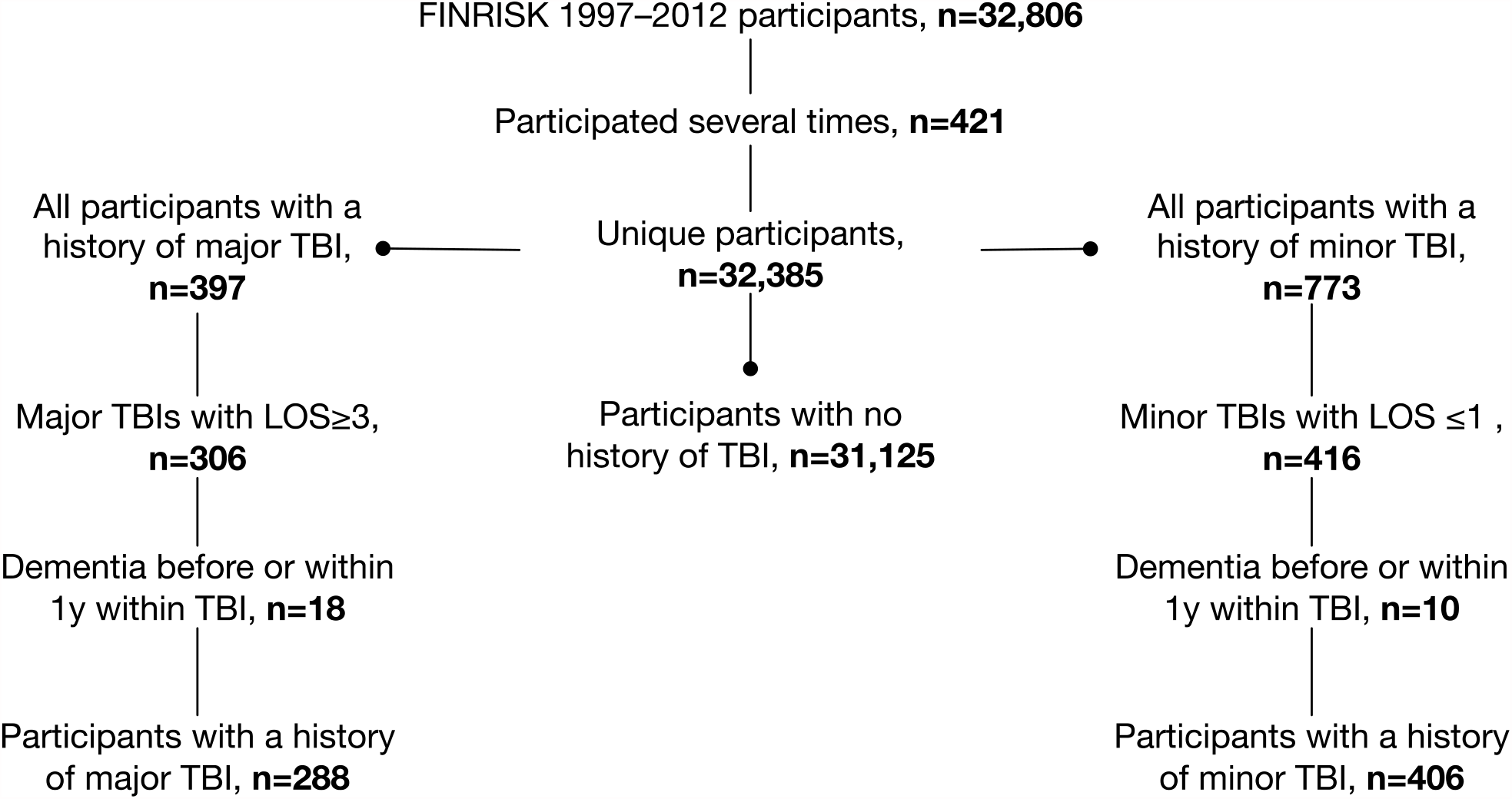
Flow chart showing the FINRISK study population according to the history of traumatic brain injury. Abbreviations: TBI = traumatic brain injury, LOS = length of hospital stay.

Low educational status, moderate to heavy alcohol consumption, smoking, less leisure time physical activity and hypertension were more frequent among participants with major TBI than among participants with minor or no TBI (Table 1).

**Table 1:**
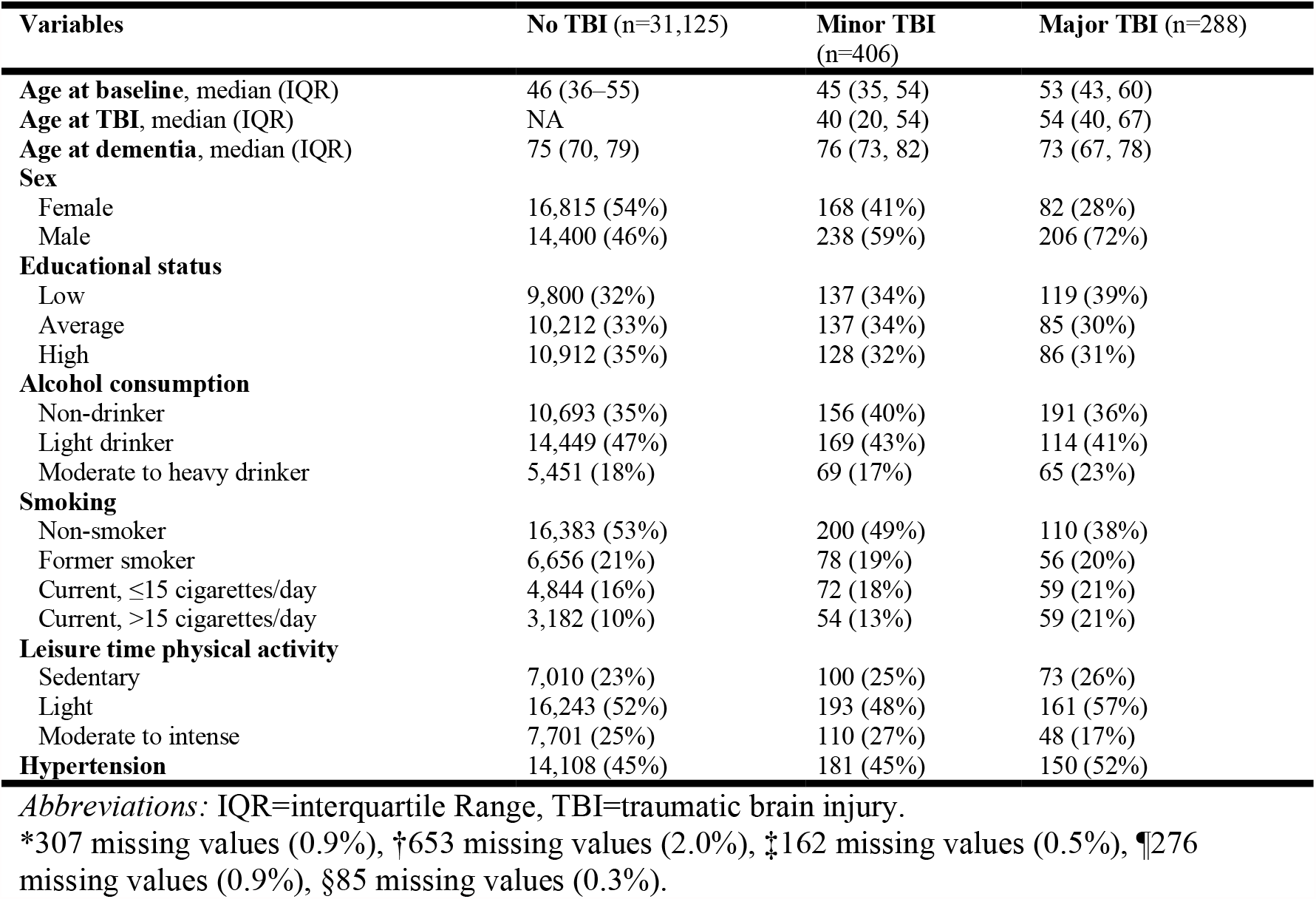
Baseline characteristics between participants with no history of traumatic brain injury, minor and major traumatic brain injury

During follow-up, 9.9% (n = 3,095) of participants with no TBI died, compared to 36.1% (n = 104) of participants with major TBI and 13.1% (n = 53) of participants with minor TBI. The median age of death for participants in the no TBI group was 69.3 years, compared to 70.6 years for participants in the major TBI group and 71.4 years for participants in the minor TBI group (Online Supplemental Table 3). Major TBI increased the risk of death, with an HR of 1.85 (95% CI 1.52–2.45) after adjusting for sex and age. In contrast, minor TBI was not associated with an increased risk of death (HR 1.17, 95% CI 0.89–1.54).

### Risk of dementia after traumatic brain injury in the FINRISK cohort

There were a total of 1,010 new dementia cases (Figure 2). The median age at diagnosis was 75.4 years, and 54.9% (n = 554) of cases were female. Of the 288 participants with major TBI, 9.4% (n = 27) developed new dementia, compared to 2.2% (n = 9) and 3.0% (n = 940) of participants with minor TBI and no TBI, respectively. Participants with major TBI were younger at the time of dementia diagnosis than those with minor or no TBI (Online Supplemental Table 3). The unadjusted incidence increased with age and was highest for those with major TBI across all age groups (Online Supplemental Table 4).

**Figure 2:**
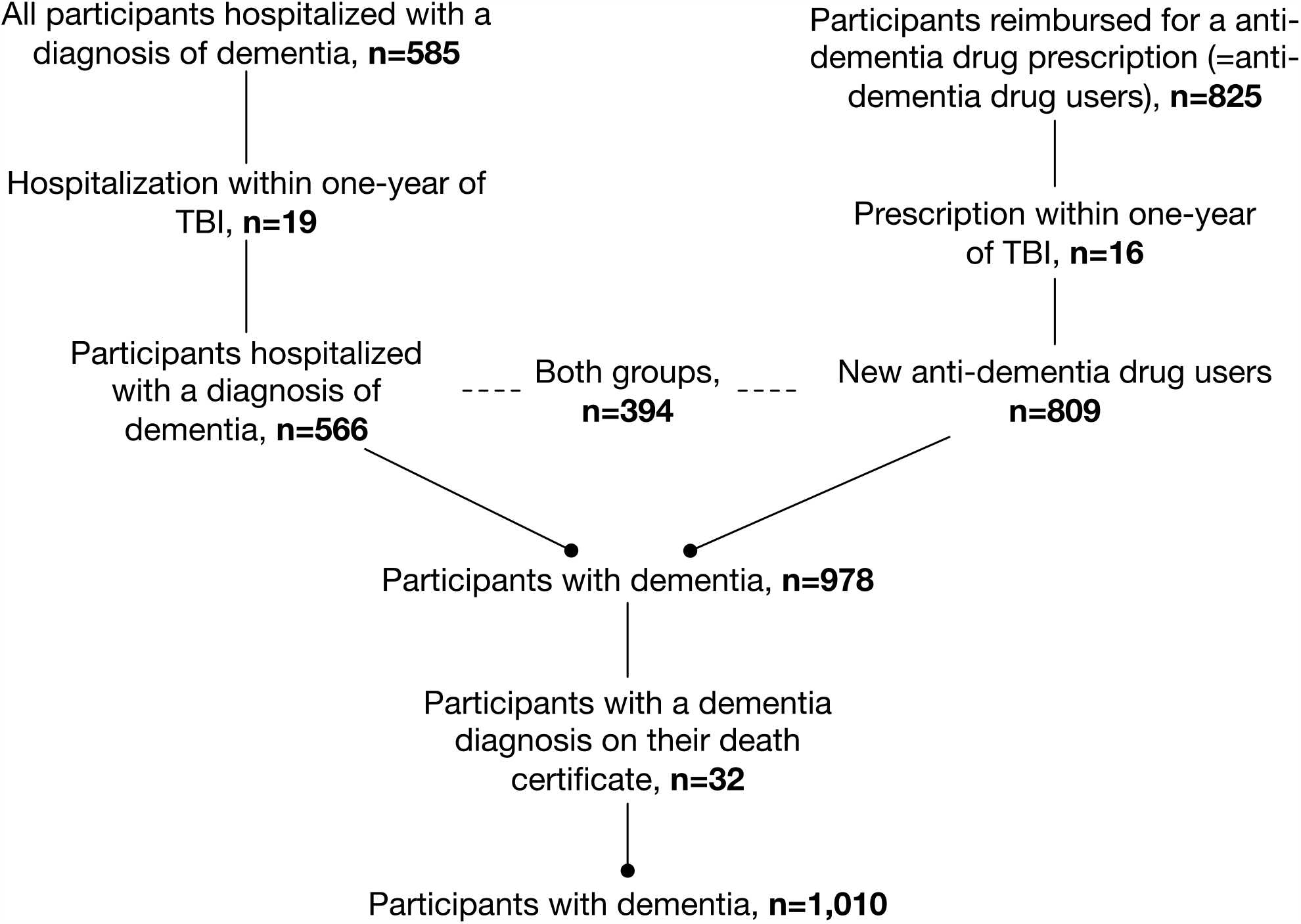
Flow chart showing dementia diagnoses in the FINRISK study population. Abbreviation: TBI = traumatic brain injury.

In the partially adjusted model, there was no association between minor TBI (HR 0.67, 95% CI 0.35– 1.29) and increased risk of dementia. Major TBI was associated with increased risk of dementia, with an HR of 1.51 (95% CI 1.03–2.22, Table 2). However, in the fully adjusted model, the association between major TBI and dementia weakened (HR 1.30, 95% CI 0.86–1.97).

**Table 2:** Hazard ratios and 95% confidence intervals from the Cox proportional hazards model by degree of adjustment

| Variable | HR (95% CI) | p-value |
| --- | --- | --- |
| **Partially adjusted model** |  |  |
| **History of TBI** |  |  |
| No TBI | 1.0 |  |
| Minor TBI | 0.67 (0.35–1.29) | 0.230 |
| Major TBI | 1.51 (1.03–2.22) | 0.036 |
| **Fully adjusted** |  |  |
| **History of TBI** |  |  |
| No TBI | 1.0 |  |
| Minor TBI | 0.64 (0.33–1.23) | 0.184 |
| Major TBI | 1.30 (0.86–1.97) | 0.219 |

Both models adjusted for adjusted for year of FINRISK study participation.
The partially adjusted model adjusts for age and sex, the fully adjusted model for all covariates (see Table 1).
*Abbreviations:* CI=confidence interval, HR=hazards ratio

In the fully adjusted risk model, higher educational status, light alcohol consumption and moderate to intensive leisure time physical activity were associated with a decreased risk of dementia (Figure 3). The risk factor analysis showed that the association between major TBI and risk of dementia was most attenuated after adjusting for alcohol consumption and leisure time physical activity (Online Supplemental Table 5).

**Figure 3:**
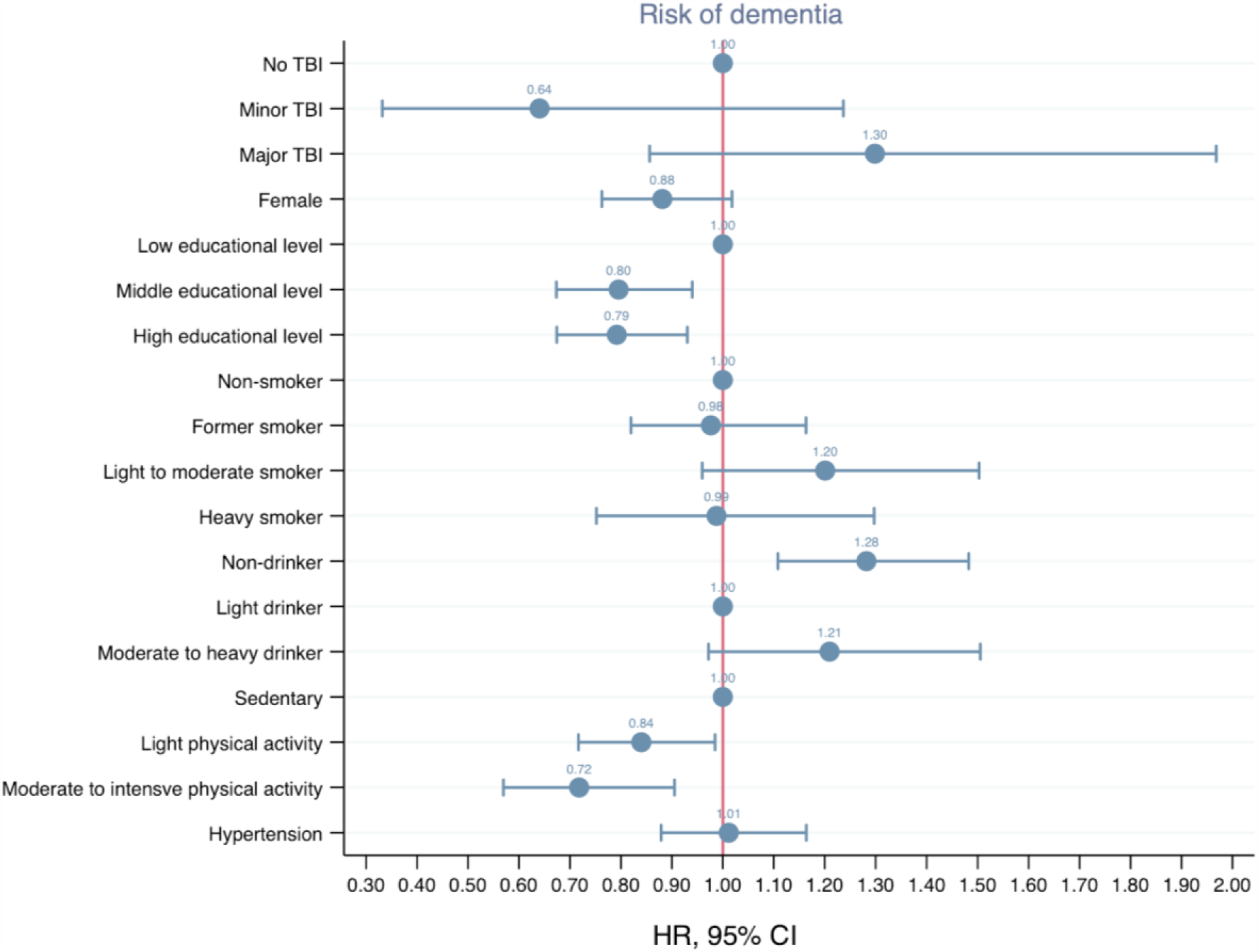
Results of the fully adjusted Cox proportional hazards regression model, showing risk factors for dementia. Abbreviations: HR = hazard ratio, CI = confidence interval, TBI = traumatic brain injury.

### Sensitivity and nested cohort analyses

According to the results of the sensitivity analysis including only those who sustained a TBI after FINRISK participation, there was no association between TBI (regardless of severity) and dementia (Online Supplemental Table 6). The predictors associated with a lower risk of dementia were the same (high education, light alcohol consumption, physical activity). The competing risks model revealed no association of either minor TBI (sHR 0.64, 95% CI 0.33–1.23) or major TBI (sHR 1.18, 95% CI 0.77–1.83) with risk of dementia (Online Supplemental Table 7).

The nested case–control analysis included 988 cases and 3,952 controls (Online Supplemental Table 8). No association between minor TBI (OR 0.72, 95% CI 0.35–1.48) or major TBI (OR 1.20, 95% CI 0.74–1.96) and dementia was found (Online Supplemental Table 9 and 10).

The nested matched exposed–nonexposed analysis included 1,477 non-TBI controls, 405 participants with minor TBI and 575 participants with major TBI (Online Supplemental Table 11). No association between minor TBI (HR 0.78, 95% CI 0.36–1.71) or major TBI (HR 1.66, 95% CI 0.90– 3.07) and dementia was found (Online Supplemental Table 12).

## DISCUSSION

### Principal findings

In this large, longitudinal FINRISK cohort, we found an association between major TBI and dementia after adjusting for age and sex, but this association weakened after adjusting for other relevant dementia risk factors (especially alcohol consumption and physical activity). We found no association between hospitalization due to minor TBI and dementia. Thus, the previously suggested association between mild TBI, and dementia appears to be the result of inadequate risk factor adjustment [2,13].

### Comparison to previous studies

The risk of dementia seems to be highest during the first year after TBI [26,27]. The proposed mechanism of TBI induced dementia relates to hyperphosphorylated tau pathology [28], persistent neuroinflammation [29] and amyloid beta pathology [30]. It is unlikely that these processes would lead to clinical manifestations of dementia within a few months of TBI [31,32]. Thus, early post-TBI diagnoses of dementia is more likely to be a result of reverse causality or due to direct TBI-related brain parenchyma injury and cognitive decline [33]. Further, the clinical manifestations of these pathological changes seem to occur only after major TBI and not after minor TBI.

TBI, especially major TBI, is associated with a high excess risk of death [7,34,35]. Thus, it is possible that some TBI survivors die before the clinical manifestation of dementia, making it seem like TBI does not increase the risk of dementia [20]. Further, TBI, dementia and early death share similar risk factors, such as low cognitive performance, less education and heavy alcohol consumption [3–5,8,9,23,24,36,37], making it essential to adjust for these when assessing the association between TBI and dementia. Based on a previous meta-analysis [13] and four new studies [26,27,38,39], TBI was added as a potential modifiable risk factor of dementia by the 2020 Lancet Commission on dementia prevention, intervention and care. However, the meta-analysis did not adjust for factors such as sex, alcohol consumption or comorbidities [13]. In addition, the four new epidemiological studies [26,27,38,39], although large in size, adjusted for comorbidities and risk factors by using hospitalized diagnostic codes (Online Supplemental File 1). This meant, for example, that physical inactivity was not accounted for in any of the four newly added studies. Further, adjusting for alcohol consumption by using register-based hospitalization diagnoses is suboptimal [40].

Insufficient risk-factor adjustment, especially in large-scale epidemiological studies, may cause misleading results [41]. A recent Atherosclerosis Risk in Communities (ARIC) study found an increased risk of dementia following head injury (HR 1.44) after adjusting for e.g. alcohol consumption, smoking, physical activity [12]. After excluding those with a diagnosis of dementia within one-year of TBI, the HR was 1.30 (95% CI 1.19–1.43), which is similar to our result for major TBI (HR 1.30, 95% CI 0.86–1.97). Differences between the ARIC study and the present study is the definition of TBI (self-reported and hospitalization in ARIC vs. hospitalization only), definition of dementia (clinical diagnosis, hospitalization and death certificate in ARIC vs. hospitalization, prescriptions and death certificate), sample size (3,440 participants with TBI and 2,350 participants with incident dementia in ARIS vs. 694 participants with TBI and 1,010 participants with incident dementia), higher age at baseline (mean 54 years in ARIC vs. median 46 years), and smaller prevalence of smoking, alcohol use and hypertension in ARIC. The differences in age and dementia-related risk factors may explain the weaker association between TBI and dementia in our cohort.

However, our results are well in line with the ARIC study, providing further evidence that major TBI is a risk factor for dementia.

### Future implications

The incidence of TBI is increasing, especially in low-and-middle income countries [1]. Although the absolute number of participants with major TBI who developed dementia was rather low in our study, approximately one in ten did develop a new diagnosis of dementia. Thus, the risk is not negligible. It is noteworthy that our and previous studies suggest that many risk factors (high alcohol consumption and physical inactivity) increase the likelihood of dementia, and these risk factors easily confound association analyses, if not properly adjusted for [9,23,42]. Indeed, TBI survivors often suffer from substance abuse and decline in cognitive performance [43]. Considering that there is no curative treatment for dementia (or TBI), secondary prevention of the effects of modifiable risk factors such as excess alcohol consumption and physical inactivity in the care and rehabilitation of TBI survivors should be prioritized.

### Strengths and limitations of the study

Several strengths of this study should be recognized. Data regarding midlife dementia risk factors were recorded prospectively through a large longitudinal study. In comparison to previous large epidemiological studies, the FINRISK study enabled us to account for self-reported lifestyle factors (e.g. alcohol consumption, smoking, physical activity) instead of relying on register-based comorbidity diagnoses. Data on hospitalized TBI and dementia were recorded prospectively (obtained through the Finnish Care Register) by the treating physician; we did not rely on the memory of participants or their relatives, minimizing the potential for recall bias. With a median follow-up time of almost 16 years per participant, we were able to study the effect of TBI on dementia in middle-aged adults when the risk of TBI is particularly high. We had virtually no loss to follow-up, making selection bias an unlikely explanation for our findings. We were able to obtain data regarding both inpatient and outpatient diagnoses of dementia at all Finnish hospitals.

Likewise, there is national and complete coverage of all patients who redeem a prescription for antidementia drugs at all Finnish pharmacies. We did not consider dementia diagnoses within one year of TBI to avoid the possibility of reverse causality. In addition, we included only ‘true’ minor TBI by selecting those who had hospitalized for no longer than one day and only ‘true’ major TBI by selecting those who were hospitalized for a minimum of three days. This reduces the possibility that participants with minor TBIs would be coded as having major TBIs and vice versa. We confirmed our results through nested cohort analyses and sensitivity analyses. Importantly, our sensitivity analysis, which used a competing risks model, strengthened our results.

Some limitations should be mentioned. Due to the limited number of participants in the FINRISK cohort who sustained a TBI, we were unable to conduct more detailed subgroup analyses (e.g. sex-specific analyses). Also, due to the limited number of persons with TBI, the failure to reject the null-hypothesis in the fully adjusted model for the association between major TBI and dementia is possibly due to lack of power (type II error). We were also unable to account for TBI-specific symptoms and findings (e.g. level of consciousness and head imaging studies). Some participants suffered from TBI before FINRISK participation, and some suffered afterwards. However, the sensitivity analysis, which included only participants who suffered from TBI after FINRISK participation, generated similar results as the primary analysis. We had no data regarding the severity of dementia. Participants with minor TBI were generally younger than participants with major TBI. Thus, it is possible that a longer follow-up (longer than 16 years) period for the minor TBI group would have been necessary to capture the association between minor TBI and dementia.

However, our nested-control analysis generated similar results as the primary analysis. Our study does not assess the association between repeat or multiple TBI and risk of dementia. The median time at which risk factor questionnaires were administered was approximately 16 years before the diagnosis of dementia. Thus, the study does not account for temporal changes in risk factor behavior. Alcohol consumption was captured through structured questionnaires, which measure only consumption the week prior to the survey. Thus, binge drinkers or former heavy drinkers who now abstain from alcohol might be included in the non-drinker group. Due to the relatively small number of participants who developed dementia, we were unable to stratify AD and non-AD dementias. Finally, the results of FINRISK participants may not be generalizable to settings outside of Finland.

## CONCLUSION

We found an association between hospitalization for at least three days or more due to major TBI and dementia. The association was attenuated after adjusting for other dementia-related risk factors, especially alcohol consumption and physical activity. There was no association between hospitalization for one day or less due to minor TBI and dementia. Earlier findings showing an association between minor TBI and dementia are possible a result of inadequate risk factor adjustment. Secondary prevention of excessive alcohol consumption and physical inactivity could decrease the risk of dementia in major TBI survivors.

## Supporting information

STROBE guidelines

## Data Availability

The datasets analyzed during the current study are not publicly available due to restrictions based in the General Data Protection Regulation (GDPR) on sensitive data such as personal health data. The access to the data may be requested through the Finnish Institute for Health and Welfare (THL) Biobank (https://thl.fi/en/web/thl-biobank/for-researchers).

## SUPPORTING INFORMATION

- **Online Supplemental File 1**. Previous studies on the association between traumatic brain injury and dementia
- **Online Supplemental File 2**. Definitions of educational status, leisure time physical activity, alcohol consumption and smoking
- **Online Supplemental Table 1**. Baseline characteristics for the whole cohort
- **Online Supplemental Table 2**. Baseline characteristics for participants with a history of minor and major traumatic brain injury before and after FINRISK participation
- **Online Supplemental Table 3**. Age at time of death and time of dementia stratified by covariates
- **Online Supplemental Table 4**. Unadjusted incidence of dementia per 10,000 person-years for participants with no traumatic brain injury and minor or major traumatic brain injury
- **Online Supplemental Table 5**. Cox proportional hazards models showing the effects of individual risk factors on the association between traumatic brain injury and risk of dementia
- **Online Supplemental Table 6**. Results of the sensitivity analysis that included only participants suffering from traumatic brain injury after FINRISK participation
- **Online Supplemental Table 7**. Results of the sensitivity analysis that used the competing risks model
- **Online Supplemental Table 8**. Characteristics of participants included in the nested case– control analysis
- **Online Supplemental Table 9**. Results of the nested case–control conditional logistic regression analysis
- **Online Supplemental Table 10**. Results of the nested case–control conditional logistic regression analysis after excluding controls who later developed dementia
- **Online Supplemental Table 11**. Characteristics of participants included in the matched exposed–nonexposed analysis
- **Online Supplemental Table 12**. Risk of dementia based on matched exposed–nonexposed Cox proportional hazards analyses

## ACKNOWLEDGEMENTS

The authors wish to thank all researchers and collaborators participating in the FINRISK project.

## FINANCIAL DISCLOSURE STATEMENT

RR has received research grants from Medicinska Understödsföreningen Liv & Hälsa, Finska Läkaresällskapet and Svenska Kulturfonden. JK has been supported by the Academy of Finland (grant 312073). The funders had no role in study design, data collection and analysis, decision to publish, or preparation of the manuscript.

## COMPETING INTERESTS

The authors declare no competing interests.

## ETHICS STATEMENT

Written consent was obtained from each participant and the surveys obtained permissions from the ethics committee, which varied over time. For the first used in this study, in 1997, the approval was obtained from the Ethics Committee for the National Public Health Institute. For the three latest surveys, in 2002, 2007 and 2012, the approval was obtained from the Ethics Committee for the Helsinki and Uusimaa Hospital District. For secondary use of the FINRISK database, the National Institute for Health and Welfare approved the study and granted us access to the database (THL/155/6.00.00/2019).

## Notes

### Competing Interest Statement

The authors have declared no competing interest.

### Funding Statement

RR has received research grants from Medicinska Understodsforeningen Liv & Halsa, Svenska Lakaresallskapet. JK has been supported by the Academy of Finland (grant 312073). The funders had no role in study design, data collection and analysis, decision to publish, or preparation of the manuscript.

