## Supplementary material for "Risk of dementia after hospitalization due to traumatic brain injury: a longitudinal, population-based study": STROBE guidelines

STROBE Statement—checklist of items that should be included in reports of observational studies

|  | Item No. | Recommendation | Page  No. | Relevant text from manuscript |
| --- | --- | --- | --- | --- |
| **Title and abstract** | 1 | (*a*) Indicate the study’s design with a commonly used term in the title or the abstract | 1 | Risk of dementia after hospitalization due to traumatic brain injury: a longitudinal, population-based study |
|  |  | (*b*) Provide in the abstract an informative and balanced summary of what was done and what was found | 2 | After additional adjustment for educational status, smoking status, alcohol consumption |
| Introduction | | | |  |
| Background/rationale | 2 | Explain the scientific background and rationale for the investigation being reported | 3 | Recently, TBI was added as a new, potentially specific modifiable… |
| Objectives | 3 | State specific objectives, including any prespecified hypotheses | 3 | Our aim was to assess the independent association between… |
| Methods | | | |  |
| Study design | 4 | Present key elements of study design early in the paper | 4 | Briefly, national FINRISK surveys have been carried out in five-year intervals since… |
| Setting | 5 | Describe the setting, locations, and relevant dates, including periods of recruitment, exposure, follow-up, and data collection | 4 | the FINRISK surveys conducted in 1992, 1997, 2002, 2007 and 2012. |
| Participants | 6 | (*a*) *Cohort study*—Give the eligibility criteria, and the sources and methods of selection of participants. Describe methods of follow-up  *Case-control study*—Give the eligibility criteria, and the sources and methods of case ascertainment and control selection. Give the rationale for the choice of cases and controls  *Cross-sectional study*—Give the eligibility criteria, and the sources and methods of selection of participants | 5, 6 | We included participants between 25 and 64 years of age  We defined minor TBI as an International Statistical...  We defined major TBI as an...  conducted a nested case-control analysis by matching participants by analysis time... |
|  |  | (*b*) *Cohort study*—For matched studies, give matching criteria and number of exposed and unexposed  *Case-control study*—For matched studies, give matching criteria and the number of controls per case |  |  |
| Variables | 7 | Clearly define all outcomes, exposures, predictors, potential confounders, and effect modifiers. Give diagnostic criteria, if applicable | 5, 6 | Educational status was defined as low, middle and high. Smoking status was defined as non-smoker...  We defined the date of dementia diagnosis as the date... |
| Data sources/ measurement | 8* | For each variable of interest, give sources of data and details of methods of assessment (measurement). Describe comparability of assessment methods if there is more than one group | 5, 6 | Educational status was defined as low, middle and high. Smoking status was defined as non-smoker...  We defined the date of dementia diagnosis as the date... |
| Bias | 9 | Describe any efforts to address potential sources of bias | 4, 5 | To reduce the likelihood of participants with minor TBI to have a major TBI we only considered  To minimize the possibility of reverse causality |
| Study size | 10 | Explain how the study size was arrived at | Figure 1 |  |

Continued on next page

| Quantitative variables | 11 | Explain how quantitative variables were handled in the analyses. If applicable, describe which groupings were chosen and why | 5, 6 | Educational status was defined as low, middle and high. Smoking status was defined as non-smoker... |
| --- | --- | --- | --- | --- |
| Statistical methods | 12 | (*a*) Describe all statistical methods, including those used to control for confounding | 6 | To assess the association between TBI and risk of dementia |
|  |  | (*b*) Describe any methods used to examine subgroups and interactions | 6 | We further conducted a nested case-control analysis |
|  |  | (*c*) Explain how missing data were addressed | 6 | Because of low number of missing data per variable, we used complete case analyses |
|  |  | (*d*) *Cohort study*—If applicable, explain how loss to follow-up was addressed  *Case-control study*—If applicable, explain how matching of cases and controls was addressed  *Cross-sectional study*—If applicable, describe analytical methods taking account of sampling strategy | 6 | e defined end of follow-up at any cause of death, date of dementia diagnosis... |
|  |  | (*e*) Describe any sensitivity analyses | 6 | we performed a sensitivity analysis, including only those being...  We further conducted a nested case-control analysis |
| Results | | | | |
| Participants | 13* | (a) Report numbers of individuals at each stage of study—eg numbers potentially eligible, examined for eligibility, confirmed eligible, included in the study, completing follow-up, and analysed | Figure 1 |  |
|  |  | (b) Give reasons for non-participation at each stage | Figure 1 |  |
|  |  | (c) Consider use of a flow diagram | Figure 1 |  |
| Descriptive data | 14* | (a) Give characteristics of study participants (eg demographic, clinical, social) and information on exposures and potential confounders | 7 | The study population constituted of 32,385 individuals |
|  |  | (b) Indicate number of participants with missing data for each variable of interest | Table 1, Online Supplemental Table 1 |  |
|  |  | (c) *Cohort study*—Summarise follow-up time (eg, average and total amount) | 7 | Total time at risk was 500,954 person-years (median 15.8 years) |
| Outcome data | 15* | *Cohort study*—Report numbers of outcome events or summary measures over time | 7, Figure 2 | There was a total of 1,010 new dementia cases (Figure 2). |
|  |  | *Case-control study—*Report numbers in each exposure category, or summary measures of exposure |  |  |
|  |  | *Cross-sectional study—*Report numbers of outcome events or summary measures |  |  |
| Main results | 16 | (*a*) Give unadjusted estimates and, if applicable, confounder-adjusted estimates and their precision (eg, 95% confidence interval). Make clear which confounders were adjusted for and why they were included | Online Supplemental Table 3, Table 2, 7, 8 | in the fully adjusted model, the association between major TB |
|  |  | (*b*) Report category boundaries when continuous variables were categorized | NA |  |
|  |  | (*c*) If relevant, consider translating estimates of relative risk into absolute risk for a meaningful time period |  |  |

Continued on next page

| Other analyses | 17 | Report other analyses done—eg analyses of subgroups and interactions, and sensitivity analyses | 8 | In the sensitivity analysis, including only those sustaining |
| --- | --- | --- | --- | --- |
| Discussion | | | | |
| Key results | 18 | Summarise key results with reference to study objectives | 9 | In this large longitudinal FINRISK cohort, we found |
| Limitations | 19 | Discuss limitations of the study, taking into account sources of potential bias or imprecision. Discuss both direction and magnitude of any potential bias | 11 | Some limitations should be mentioned. Due to the limited |
| Interpretation | 20 | Give a cautious overall interpretation of results considering objectives, limitations, multiplicity of analyses, results from similar studies, and other relevant evidence | 11 | Some limitations should be mentioned. Due to the limited |
| Generalisability | 21 | Discuss the generalisability (external validity) of the study results | 11 | ...FINRISK participants may not be generalizable to settings outside of Finland |
| Other information | |  | | |
| Funding | 22 | Give the source of funding and the role of the funders for the present study and, if applicable, for the original study on which the present article is based | 18 | Funding |

*Give information separately for cases and controls in case-control studies and, if applicable, for exposed and unexposed groups in cohort and cross-sectional studies.

**Note:** An Explanation and Elaboration article discusses each checklist item and gives methodological background and published examples of transparent reporting. The STROBE checklist is best used in conjunction with this article (freely available on the Web sites of PLoS Medicine at http://www.plosmedicine.org/, Annals of Internal Medicine at http://www.annals.org/, and Epidemiology at http://www.epidem.com/). Information on the STROBE Initiative is available at www.strobe-statement.org.
