## Supplementary File for "Risk of dementia after hospitalization due to traumatic brain injury: a longitudinal, population-based study"

### **Online Supplemental File 1.**

#### **Previous studies on the association between traumatic brain injury and dementia**

| **Study** | **Setting** | **Study period** | **Study sample** | **Definition of TBI** | **Definition of dementia** | **Covariate definition and adjustment** | **Follow-up time** | **Main finding** |
| --- | --- | --- | --- | --- | --- | --- | --- | --- |
| **Studies that found an association between traumatic brain injury and dementia** | | | | | | | | |
| Fann 2018*[1] | Denmark | 1977–2013 (TBI) and 1999–2013 (dementia) | Total cohort, n=2,794,852  TBI, n=132,093, Dementia, n=126,734  Persons aged ≥50 years on January 1, 1995 were considered | Register based ICD-8/ICD-10 | ICD-8/ICD-10 and/or at least one anti-dementia drug prescription | Age, sex, marital status, ICD-8/ICD-10 codes for medical comorbidities, neurological comorbidities, psychiatric comorbidities, drug-related abuse, alcohol-related abuse | Mean 9.9 years per patient | TBI versus no TBI increased the risk of dementia by a HR of 1.24–1.58  Mild TBI increased the risk of dementia by a HR of 1.17–1.46  Severe TBI increased the risk of dementia by a HR of 1.35–1.91 |
| Nordström 2018*[2] | Sweden | 1964–2012 (TBI and dementia) | Cohort I, n=164,334 TBI and controls, Cohort II; n=136,233 dementia and controls; Cohort III, n=46,970 sibling pairs discordant for TBI  Persons aged ≥50 years on December 31, 2005 were considered | Register based ICD-8/ICD-9/ICD-10 | Register based ICD-8/ICD-9/ICD-10 | Age, sex, civil status, education, early retirement pension, ICD-8/ICD-9/ICD-10 codes for medical comorbidities, alcohol intoxication and depression | Cohort I: mean 15.3 years, Cohort II: mean 18.8 years | TBI versus no TBI increased the risk of dementia by an OR of 1.71–1.89 |
| Yaffe 2019*[3] | U.S. Veterans (women) | 2004–2015 | n=109,140 females ≥55 years | Register based ICD-9 | Register based ICD-9 | ICD-9 for medical and psychiatric comorbidities, demographics (age, race/ethnicity, ZIP codes for socioeconomic classification) | Mean 4.0 years | TBI increased risk the risk of dementia by an sHR of 1.5 (95% CI 1.0–2.2) |
| Chu 2016*[4] | Taiwan | 2004–2005 | n=12,931 TBI and n=51,724 controls | Register based ICD-9 | Register based ICD-9 | Age, sex, urbanization level, income, ICD-9 for comorbidities | Median 2.0 years | TBI increased the risk of dementia by a HR of 3.2 (95% CI 2.7–3.9) |
| Luukinen 2005**[5] | Finland | 1991–2000 | n=152 persons ≥70 years | Hospitalization to university hospital | DSM-IV | Age, sex, education, diabetes, physical disability, alcohol use (self-reported), diabetes, BMI, MMSE, APOE 𝝴4 | 9 years | TBI increased the risk of dementia by a HR of 2.6 (95% CI 1.3–5.5) |
| Sundström 2007**[6] | Sweden | 1988–1990, 1993–1995, 1998–2000 | n=543 | Self-reported and validated | DSM-IV | Age, sex, APOE 𝝴4 | At least 5 years | TBI without APOE 𝝴4 did not increase the risk for dementia (OR 0.9, 95% CI 0.4–1.8), but TBI with APOE 𝝴4 increased the risk for dementia (OR 5.2, 95% CI 2.0–14.0) |
| Luukinen 2008**[7] | Finland | 1991–1992 | n=123 persons ≥70 years | Self-reported and validated | DSM-IV | Age, MMSE score, sex, educational status, APOE 𝝴4 | 9 years | TBI without APOE 𝝴4 did not increase the risk for dementia (OR 1.5, 95% CI 0.5–4.4), but TBI with APOE 𝝴4 increased the risk for dementia (OR 2.7, 95% CI 1.02–7.2) |
| Abner 2014**[8] | United States | 1989–2004 | n=649 ≥60 years | Self-reported | Post-mortem neuropathological diagnosis | Age, sex, education, APOE 𝝴4, family history of dementia, hypertension and smoking (self-reported) | Not specified | TBI increased the risk for pathological diagnosis of AD by an OR of 1.5 in men but not in women |
| Rasmusson 1995**[9] | United States | Not specified | n=68 persons with dementia and n=34 spouse controls | Self-reported by spouse informants | NINCDS-ADRDA Alzheimer's Criteria and neuropathological findings | Age, sex, education | Not specified | 20% of familial and 43.5% of sporadic AD had premorbid history of TBI (OR 3.1). TBI had no effect of age of dementia onset |
| Nemetz 1999**[10] | United States | 1935–1984 (TBI) | n=1,283 persons with TBI | Manual review of medical records | Manual review of medical records | Age, sex | Until 1988 | History of TBI shortened the time to AD diagnosis (median 10 years vs. 18 years from TBI, p=0.015). Calculated as the difference between observed and expected time of AD diagnosis. |
| Barnes 2014**[11] | United States | 2000–2003 (baseline) and 2003–2012 (follow-up) | n=188,764 veterans ≥ years at baseline | Register based ICD-9 | Register based ICD-9 | Age, sex, ZIP code for education and income, ICD-9 codes for medical and psychiatric comorbidities | Mean 7.4 years | TBI increased the risk for dementia by a sHR of 1.8 (95% CI 1.6–2.1) |
| Gardner 2014**[12] | California, US | 2005–2011  2005–2006 (TBI) | n=51,799 persons with TBI ≥55 years | Register based ICD-9 | Register based ICD-9 | Age, sex, race, ICD-9 codes for medical comorbidities, trauma mechanism, trauma severity, health care use, ZIP code for socioeconomic classification | Median follow-up 6 years  No censoring at death | TBI increased the risk of dementia by a HR of 1.3 (95% CI 1.2–1.3)  Mild TBI increased the risk for dementia in persons ≥65 years (HR 1.5, 95% CI 1.3–1.6) but not in persons 55–64 years)  Moderate-to-severe TBI increased the risk for TBI in all persons ≥55 years (for those 55-64 years HR 1.7, 95% CI 1.4–2.1, for those 65-74 years HR 1.5, 95% CI 1.3–1.6) |
| Nordström 2014**[13] | Sweden | 1969–1986 | n=811,622 men | Register based ICD-8/ICD-9/ICD-10 | Register based ICD-8/ICD-9/ICD-10 | Age, place and year of conscription, weight, height, knee strength, blood pressure, cognitive function score, education, income, heritability (dementia and TBI in parents), ICD-8/ICD-9/ICD-10 diagnoses during follow up (including alcohol intoxication, drug intoxication, depression, stroke | Median 33 years | One mild TBI increased the risk for dementia by a HR of 1.5 (95% CI 1.1–2.0). Specifically, one mild TBI increased the risk for non-AD dementia (HR 1.7, 95% CI 1.2–2.3) but not for AD (HR 1.0, 95% CI 0.5–2.0)  One severe TBI increased the risk for dementia by a HR of 2.3 (95% CI 1.5–3.6). Specifically, one severe TBI increased the risk for non-AD dementia (HR 2.6, 95% CI 1.6–4.1) but not for AD (HR 0.7, 95% CI 0.1–5.2) |
| Wang 2012**[14] | Taiwan | 2000–2004 (TBI) | n=44,925 persons with TBI and n=224,625 controls without TBI | Register based ICD-9 | Register based ICD-9 | Age, sex, area of living, ICD-9 codes for comorbidities | 5-year follow-up | TBI increased the risk of dementia by a HR of 1.7 (95% CI 1.6–1.8) |
| Schneider 2021[15] | United States | 1987–1989 | n=3440 persons with head injury and n=10,936 persons with no head injury | Self-reported, hospitalization data (ICD-9/ICD-10) | Cognitive assessments, telephone interview, hospitalization ICD-9 codes, death certificate | Age, race, education, family income, physical activity index, cigarette smoking, alcohol consumption, hypertension, diabetes, coronary heart disease, apolipoprotein E, depression, post-traumatic stress disorder | Median 25 years | Head injury increased the risk of dementia by a HR of 1.44 (95% CI 1.32–1.57). In a model adjusting for competing risk, HR was 1.72 (95% CI 1.57–1.89). The risk was higher among women than among men. |
| Tolppanen 2017[16] | Finland | 2005–2011 (diagnosis of AD)  1972–2011 (TBI) | n=70,719 persons with AD and 282,862 controls | Register based ICD-9/ICD-10 | Clinically verified diagnosis of AD | Age, sex, psychotropic and epileptic drugs (ATC codes), comorbidities and substance abuse (ATC codes and ICD-10), socioeconomic status | Not stated | TBI versus no TBI increased the risk of AD by an OR of 1.23 (95% CI 1.18–1.29)  Mild TBI increased the risk of AD by an OR of 1.19 (95% CI 1.12–1.25)  Severe TBI increased the risk of AD by an OR of 1.36 (95% CI 1.25–1.48) |
| Barnes 2018[17] | U.S. Veterans (91% men) | 2001–2014 (TBI) | n=178,779 persons with TBI and 178,779 controls | Clinical evaluation and ICD-9 | Register based ICD-9 | ICD-9 for medical comorbidities, psychiatric disorders and substance abuse, self-reported demographic information (age, sex, race/ethnicity) | Not reported | Mild TBI without loss of consciousness increased the risk of dementia by an HR of 2.4 (95% CI 2.1–2.7)  Mild TBI with loss of consciousness increased the risk of dementia by an HR of 2.5 (95% CI 2.3–2.8)  Moderate-to-severe TBI increased the risk of dementia by an HR of 3.8 (95% CI 3.6–3.9) |
| Redelmeier 2019[18] | Ontario, Canada | 1993–2013 (TBI) | n=28,815 persons ≥66 years with concussion | Register based ICD-9 | Register based ICD-9 | Age, sex, socioeconomic quintile, home location, Ontario Drug Benefit program database for use of cardiovascular medications, neuropsychiatric medications, and miscellaneous medications | Mean 3.9 years | One in six persons with concussion developed dementia during a mean follow-up of 3.9 years |
| Lee 2013[19] | Taiwan | 2005–2009 | n=28551 persons with mild TBI | Register based ICD-9 | ICD-9 and anti-dementia drug prescription or a "catastrophic illness certificate" application | Age, gender, urbanization level, socioeconomic status, ICD-9 codes for medical comorbidities | 1 year | Mild TBI increased risk for dementia by a HR of 3.3 (95% CI 2.7–3.9). Median time from mild TBI to dementia was 1.0 years (95% CI 0.8–1.2) |
| Raj 2017[20] | Finland | 1987–2014 | n=19,936 persons with moderate-to-severe TBI and n=20,7033 persons with mild TBI | Register based ICD-9/ICD-10 | Register based ICD-9/ICD-10 | Age, sex, socioeconomic status, education level | Mean 11 years | Moderate-to-severe TBI increased the risk of dementia compared to mild TBI with a HR of 1.9 (95% CI 1.6–2.2) |
| Guo 2000[21] | United States, Canada, and Germany | 1991–1996 | n=2,233 persons with AD and unaffected spouse(s) controls | Informant questionnaire | NINCDS-ADRDA Alzheimer's Criteria | Age and sex | NA | Head injury with loss of consciousness increased the risk of dementia by an OR of 9.9 (95% CI 6.5–15.1)  Head injury without loss of consciousness increased the risk of dementia by an OR of 3.1 (95% CI 2.3–4.0) |
| **Studies that did not find any association between traumatic brain injury and dementia** | | | | | | | | |
| Mehta 1999**[22] | Netherlands (Rotterdam) | 1990–1993 | n=6,645 persons ≥55 years | Self-reported questionnaire | Dementia diagnosis according to the DSM-III-R criteria  AD diagnosis according to NINCDS-ADRDA Alzheimer's Criteria | Age, sex, education | Mean 2.1 years | History of head trauma did not significantly increase the risk of (loss of consciousness, multiple TBIs, duration of loss of consciousness did not affect) |
| Dams-O'Connor 2013[23] | United States | 1994–2010 | n=3,466 persons ≥65 years | Self-reported questionnaire | Dementia diagnosis according to the DSM-IV criteria  AD diagnosis according to NINCDS-ADRDA Alzheimer's Criteria | Age, sex, education, APOE 𝝴4 | Mean 7.4 years | History of TBI with loss of consciousness did not significantly increase risk for dementia independent of age at the time of head injury |
| Crane 2016[24] | United States | 1994–2004 | Cohort I, n=3,666  Cohort II, n=2,689 | Self-reported questionnaire | Dementia diagnosis according to the DSM-IV criteria  AD diagnosis according to NINCDS-ADRDA Alzheimer's Criteria  Clinical, neuropsychological and cognitive testing | Age, sex, education, APOE 𝝴4 | Cohort I: Mean 7.8 years  Cohort II: Mean 5.5 years | History of TBI with loss of consciousness did not significantly increase risk for dementia |
| *New added evidence included in 2020 Lancet Commission on dementia prevention, intervention and care.  **Studies included as original evidence used by 2020 Lancet Commission on dementia prevention, intervention and care stemming from a meta-analysis by Huang and colleagues [25]  AD=Alzheimer's Disease, BMI=Body Mass Index, CI=Confidence Interval, HR=Hazards Ratio, MMSE=Mini Mental State Examination, OR=Odds Ratio, sHR=sub-Hazard Ratio, NINCDS-ADRDA=National Institute of Neurological and Communicative Disorders and Stroke/Alzheimer’s Disease and Related Disorders Association, DSM-=Diagnostic and Statistical Manual of Mental Disorders | | | | | | | | |

### **Online Supplemental File 2.**

#### **Educational status definition**

Educational status was classified as low, middle and high. Educational status was calculated by dividing years of formal education into tertiles within each birth cohort, separately for men and women. This in order to account for the steep increase in rise in schooling during the 20th century.[26]

#### **Leisure time physical activity definition**

Leisure time physical activity was defined as follows:

1. Sedentary: In my leisure time I read, watch TV, and work in the household with tasks which do not make me move much and which do not physically tax me.
2. Light: In my spare time I walk, cycle or exercise otherwise at least 4 hours per week. This includes walking, fishing and hunting, light gardening etc. but excludes travel to work.
3. Moderate: In my spare time I exercise to maintain my physical condition, e.g. running, jogging, skiing, gymnastics, swimming, playing ball games or I do heavy gardening or the like for at least 3 hours per week.
4. Intensive: In my spare time I regularly exercise competitive-wise several times a week running, orienteering, skiing, swimming, playing ball games or other heavy sports.

The classification has shown good criterion validity against morbidity and mortality and moderate correlation against accelometer counts among the working-age population.[27] The categories moderate activity and 4 (intensive activity were combined due to the low number of participants in the intensive group (<2% of all).

#### **Alcohol consumption definition**

Alcohol consumption was assessed with structured questionnaires regarding the average amount of alcohol they had consumed the week prior to the survey. Average alcohol intake (grams per day) was calculated as the sum of the daily number of drinks multiplied by the average alcohol content per type of alcoholic beverage.

The amount of ethanol in different beverages was quantitated based on defined portion sizes as follows: regular beer 12 grams (1/3 L), strong beer 15.5 grams (1/3 L), long drink 15.5 grams (1/3 L), spirit 12 grams (4 cL), wine 12 grams (12 cL) and cider 12 grams (1/3 L). A dose of 12 grams of pure ethanol was considered as one standard drink.

The data on alcohol consumption was subsequently used to categorize the population by gender and drinking habits as follows: i) participants who reported no current alcohol consumption were referred to as non-drinkers, ii) light drinkers consumed between 1–13 drinks (men) or 1–6 drinks (women), iii) moderate drinkers consumed 14–23 drinks (men) or 7–15 drinks (women) and iv) heavy drinkers consumed more than 23 drinks (men) or more than 15 drinks (women) per week.[28]

For the analyses, moderate and heavy drinkers were combined into one category due to the low number of participants in the heavy drinking group (<5%).

#### **Smoking definition**

Current smoker (including those that stopped less than 6 months ago), former smoker (stopped more than 6 months ago), and non-smoker. Current regular smokers were divided current smokers based upon number of cigarettes used to ≤15 cigarettes per day (light to moderate smokers) and >15 cigarettes per day (heavy smokers).[29] The non-smoker group may contain occasional smokers.

### **Online Supplemental Table 1.** Baseline characteristics for the whole cohort

| **Variables** | **All participants**  (n=32,385) | **Men**  (n=15,106) | **Women**  (n=17,279) |
| --- | --- | --- | --- |
| **Age at baseline**, median (IQR) | 46 (36, 55) | 46 (36, 56) | 45 (35, 55) |
| **Sex** |  |  |  |
| Female | 17,279 (53%) | NA | 17,279 (100%) |
| Male | 15,106 (47%) | 15,106 (100%) | NA |
| **Educational status*** |  |  |  |
| Low | 10,206 (32%) | 4,663 (31%) | 5,543 (32%) |
| Average | 10,599 (33%) | 4,926 (33%) | 5,673 (33%) |
| High | 11,273 (35%) | 5,367 (36%) | 5,906 (34%) |
| **Alcohol consumption**† |  |  |  |
| Non-drinker | 11,120 (35%) | 3,925 (27%) | 7,195 (42%) |
| Light drinker | 14,928 (47%) | 7,843 (53%) | 7,085 (42%) |
| Moderate to heavy drinker | 5,683 (18%) | 2,981 (20%) | 2,712 (16%) |
| **Smoking**‡ |  |  |  |
| Non-smoker | 16,919 (53%) | 6,322 (42%) | 10,597 (62%) |
| Former smoker | 6,897 (21%) | 3,920 (26%) | 2,977 (17%) |
| Current, ≤15 cigarettes/day | 5,048 (16%) | 2,3332 (16%) | 2,716 (16%) |
| Current, >15 cigarettes/day | 3,359 (10%) | 2,449 (16%) | 910 (5%) |
| **Leisure time physical activity**¶ |  |  |  |
| Sedentary | 7,302 (23%) | 3,359 (23%) | 3,943 (23%) |
| Light | 16,834 (52%) | 7,526 (50%) | 9,308 (54%) |
| Moderate to intense | 7,973 (25%) | 4,103 (27%) | 3,870 (23%) |
| **Hypertension**§ | 14,669 (45%) | 7,337 (48%) | 7,332 (43%) |
| *Abbreviations: IQR=interquartile range.*  *307 missing values (0.9%), †653 missing values (2.0%), ‡162 missing values (0.5%), ¶276 missing values (0.9%), §85 missing values (0.3%) | | | |

### **Online Supplemental Table 2.** Baseline characteristics for participants with a history of minor and major traumatic brain injury before and after FINRISK participation

| **Variable** | **Minor TBI before entry** (n=238) | **Minor TBI after entry** (n=168) | **p-value** | **Major TBI before entry** (n=127) | **Major TBI after entry** (n=161) | **p-value** |
| --- | --- | --- | --- | --- | --- | --- |
| **Age at baseline**, median (IQR) | 43 (32, 53) | 47 (39, 55) | 0.007 | 52 (41, 59) | 55 (45, 61) | 0.053 |
| **Age at TBI**, median (IQR) | 24 (16, 39) | 56 (47, 67) | <0.001 | 40 (23, 50) | 64 (55, 74) | <0.001 |
| **Age at dementia**, median (IQR) | 76 (69, 80) | 78 (72, 84) | 0.807 | 73 (66, 77) | 73 (69, 80) | 0.651 |
| **Sex** |  |  |  |  |  |  |
| Female | 99 (42%) | 69 (41%) | 0.916 | 35 (28%) | 47 (29%) | 0.760 |
| Male | 139 (58%) | 99 (59%) |  | 92 (72%) | 114 (71%) |  |
| **Educational status** |  |  |  |  |  |  |
| Low | 74 (31%) | 63 (38%) | 0.227 | 53 (43%) | 57 (37%) | 0.535 |
| Average | 88 (37%) | 49 (30%) |  | 36 (29%) | 49 (31%) |  |
| High | 75 (32%) | 53 (32%) |  | 35 (28%) | 51 (32%) |  |
| **Alcohol consumption** |  |  |  |  |  |  |
| Non-drinker | 87 (38%) | 69 (43%) | 0.570 | 52 (42%) | 49 (31%) | 0.178 |
| Light drinker | 104 (44%) | 65 (40%) |  | 47 (38%) | 67 (43%) |  |
| Moderate to heavy drinker | 41 (18%) | 28 (17%) |  | 25 (20%) | 40 (26%) |  |
| **Smoking** |  |  |  |  |  |  |
| Non-smoker | 114 (48%) | 86 (52%) | 0.594 | 39 (31%) | 71 (44%) | 0.113 |
| Former smoker | 51 (21%) | 27 (16%) |  | 26 (21%) | 30 (19%) |  |
| Current, ≤15 cigarettes/day | 42 (18%) | 30 (18%) |  | 32 (26%) | 27 (17%) |  |
| Current, >15 cigarettes/day | 30 (13%) | 24 (14%) |  | 27 (22%) | 32 (20%) |  |
| **Leisure time physical activity** |  |  |  |  |  |  |
| Sedentary | 54 (23%) | 46 (28%) | 0.107 | 41 (34%) | 32 (20%) | 0.035 |
| Light | 110 (46%) | 83 (50%) |  | 62 (50%) | 99 (62%) |  |
| Moderate to intense | 74 (31%) | 36 (22%) |  | 19 (16%) | 29 (18%) |  |
| **Hypertension** | 108 (45%) | 73 (43%) | 0.701 | 55 (43%) | 95 (59%) | 0.008 |
| *Abbreviations: IQR=interquartile range, TBI=traumatic brain injury.* | | | | | | |

### **Online Supplemental Table 3.** Age at time of death and time of dementia stratified by covariates

| **Variables** | **Median age at time of death (IQR)**  Total N = 3,339 | | **Median age at time of dementia (IQR)**  Total N = 1,010 |
| --- | --- | --- | --- |
| **All** | 69.3 (61.5–76.6) |  | 75.4 (70.2–79.5) |
| **No TBI** | 69.3 (61.6–76.6) |  | 75.5 (70.2–79.5) |
| **Minor TBI** | 71.4 (58.9–77.7) |  | 76.4 (72.9–82.1) |
| **Major TBI** | 70.6 (60.8–76.1) |  | 72.7 (67.1–78.1) |
| **Sex** |  |  |  |
| Female | 71.1 (62.7–78.3) | | 74.7 (69.5–79.3) |
| Male | 68.3 (60.7–75.4) | | 75.7 (70.5–79.8) |
| **Educational status** |  |  |  |
| Low | 68.1 (60.7–75.2) | | 74.3 (70.0–78.9) |
| Average | 69.0 (60.9–76.6) | | 75.7 (70.0–79.8) |
| High | 70.9 (63.0–77.9) | | 76.0 (70.8–79.6) |
| **Alcohol consumption** |  |  |  |
| Non-drinker | 71.4 (63.9–78.1) | | 75.7 (70.6–79.8) |
| Light drinker | 69.5 (61.6–76.7) | | 75.5 (70.2–79.7) |
| Moderate to heavy drinker | 65.2 (57.6–72.6) | | 72.5 (66.6–77.5) |
| **Smoking** |  |  |  |
| Non-smoker | 72.2 (63.4–79.1) | | 76.1 (71.0–80.1) |
| Former smoker | 71.4 (64.7–77.4) | | 75.4 (69.7–79.2) |
| Current, ≤15 cigarettes/day | 66.2 (57.8–73.6) | | 73.3 (68.0–77.6) |
| Current, >15 cigarettes/day | 65.4 (58.3–71.7) | | 70.6 (64.8–77.4) |
| **Leisure time physical activity** |  |  |  |
| Sedentary | 67.1 (59.9–74.8) | | 73.2 (67.9–78.5) |
| Light | 70.5 (62.9–77.3) | | 75.9 (71.0–80.0) |
| Moderate to intense | 68.3 (57.8–76.3) | | 75.2 (69.2–79.5) |
| **Hypertension** |  | |  |
| No | 66.0 (57.6–74.5) | | 74.4 (69.4–79.0) |
| Yes | 70.5 (63.5–77.4) | | 75.6 (70.5–79.8) |
| *Abbreviations: IQR=interquartile range.* | | | |

### **Online Supplemental Table 4.** Unadjusted incidence of dementia per 10,000 person-years for participants with no traumatic brain injury and minor or major traumatic brain injury

| **Variable** | **Unadjusted incidence of dementia per 10,000 person-years (95% CI)** | | |
| --- | --- | --- | --- |
| **Age at baseline** | **No TBI** (n=31,125) | **Minor TBI** (n=406) | **Major TBI** (n=288) |
| **25–39 years** | 0.5 (0.2–0.9) | NA* | 10.3 (1.4–72.9) |
| **40–49 years** | 6.3 (5.1–7.8) | NA* | 36.3 (13.6–96.8) |
| **50–59 years** | 23.1 (26.2–32.5) | 13.6 (3.4–54.5) | 70.0 (37.7–130.2) |
| **60–64 years** | 86.0 (78.6–94.2) | 91.5 (43.6–191.8) | 109.5 (62.2–192.8) |
| *Abbreviations*: CI=confidence intervals, TBI=traumatic brain injury.  *No dementia cases | | | |

### **Online Supplemental Table 5.** Cox proportional hazards models showing the effects of individual risk factors on the association between traumatic brain injury and risk of dementia

|  | **Hazard ratio (95% confidence intervals)** | | | | | |
| --- | --- | --- | --- | --- | --- | --- |
| **Category** | **No risk factors** | **Educational level** | **Smoking** | **Alcohol consumption** | **Physical activity** | **Hypertension** |
| **No TBI** | 1.0 | 1.0 | 1.0 | 1.0 | 1.0 | 1.0 |
| **Minor TBI** | 0.67 (0.35–1.29) | 0.65 (0.34–1.26) | 0.67 (0.35–1.29) | 0.66 (0.34–1.27) | 0.66 (0.34–1.27) | 0.67 (0.35–1.29) |
| **Major TBI** | 1.51 (1.03–2.22) | 1.48 (1.00–2.18) | 1.47 (0.99–2.18) | 1.39 (0.93–2.07) | 1.41 (0.94–2.10) | 1.51 (1.03–2.21) |
| *all models adjusted for sex, year of FINRISK participation  Education level categories are low, middle and high.  Smoking categories are non-smoker, former smoker, current smoker ≤15 cigarettes per day, current smoker >15 cigarettes per day.  Alcohol consumption are categorized according to gender and number of drinks per day (*please see "Alcohol consumption definition")*  Leisure time activity categories are sedentary, light and moderate to intense.  Hypertension categories are no hypertension and hypertension. | | | | | | |

### **Online Supplemental Table 6.** Results of the sensitivity analysis that included only participants suffering from traumatic brain injury after FINRISK participation

| **Variable** | **HR (95% CI)** | **p-value** |
| --- | --- | --- |
| **Partially adjusted model** |  |  |
| **History of TBI** |  |  |
| No TBI | 1.0 |  |
| Minor TBI | 0.41 (0.14–1.10) | 0.077 |
| Major TBI | 0.75 (0.40–1.41) | 0.374 |
| **Sex** |  |  |
| Male | 1.0 |  |
| Female | 0.97 (0.85–1.10) | 0.625 |
| **Fully adjusted** |  |  |
| **History of TBI** |  |  |
| No TBI | 1.0 |  |
| Minor TBI | 0.40 (0.15–1.08) | 0.070 |
| Major TBI | 0.68 (0.35–1.13) | 0.250 |
| **Sex** |  |  |
| Male | 1.0 |  |
| Female | 0.89 (0.77–1.03) | 0.111 |
| **Education status** |  |  |
| Low | 1.0 |  |
| Middle | 0.80 (0.67–0.94) | 0.009 |
| High | 0.79 (0.67–0.93) | 0.005 |
| **Smoking** |  |  |
| Non-smoker | 1.0 |  |
| Former smoker | 0.97 (0.81–1.16) | 0.728 |
| Current, ≤15 cigarettes/day | 1.21 (0.97–1.52) | 0.096 |
| Current, >15 cigarettes/day | 0.95 (0.72–1.26) | 0.724 |
| **Alcohol consumption** |  |  |
| Non-drinker | 1.27 (1.10–1.48) | 0.001 |
| Light drinker | 1.0 |  |
| Moderate to heavy drinker | 1.22 (0.97–1.52) | 0.083 |
| **Leisure time physical activity** |  |  |
| Sedentary | 1.0 |  |
| Light | 0.86 (0.73–1.01) | 0.060 |
| Moderate to intense | 0.74 (0.58–0.93) | 0.010 |
| **Hypertension** | 1.01 (0.88–1.17) | 0.853 |
| Both models adjusted for adjusted for year of FINRISK study participation  *Abbreviations*: CI=confidence interval, HR=hazards ratio | | |

### **Online Supplemental Table 7.** Results of the sensitivity analysis that used the competing risks model

| **Variable** | **sHR (95% CI)** | **p-value** |
| --- | --- | --- |
| **History of TBI** |  |  |
| No TBI | 1.0 |  |
| Minor TBI | 0.64 (0.33–1.23) | 0.178 |
| Major TBI | 1.18 (0.77–1.83) | 0.446 |
| **Education status** |  |  |
| Low | 1.0 |  |
| Middle | 0.82 (0.70–0.97) | 0.020 |
| High | 0.85 (0.73–1.00) | 0.053 |
| **Smoking** |  |  |
| Non-smoker | 1.0 |  |
| Former smoker | 0.92 (0.78–1.09) | 0.336 |
| Current, ≤15 cigarettes/day | 0.95 (0.76–1.19) | 0.660 |
| Current, >15 cigarettes/day | 0.63 (0.48–0.83) | 0.001 |
| **Alcohol consumption** |  |  |
| Non-drinker | 1.24 (1.08–1.44) | 0.003 |
| Light drinker | 1.0 |  |
| Moderate to heavy drinker | 1.04 (0.84–1.29) | 0.725 |
| **Leisure time physical activity** |  |  |
| Sedentary | 1.0 |  |
| Light | 0.92 (0.78–1.07) | 0.278 |
| Moderate to intense | 0.80 (0.64–1.01) | 0.063 |
| **Hypertension** | 1.02 (0.88–1.17) | 0.823 |
| *Abbreviations*: CI=confidence interval, sHR=subhazard ratio  Model also adjusted for year of FINRISK participation | | |

### **Online Supplemental Table 8.** Characteristics of participants included in the nested case–control analysis

| **Variables** | **Controls**  (n=3,952) | **Cases**  (n=988) |
| --- | --- | --- |
| **Age at entry**, median (IQR) | 60 (56, 63) | 60 (56, 62) |
| **Age at exit**, median (IQR) | 80 (76, 84) | 76 (70, 80) |
| **Minor TBI** | 53 (1.4%) | 9 (0.9%) |
| **Major TBI** | 81 (2.1%) | 27 (2.8%) |
| **Sex** |  |  |
| Female | 2,172 (55%) | 543 (55%) |
| Male | 1,780 (45%) | 445 (45%) |
| **Educational status*** |  |  |
| Low | 1,094 (28%) | 317 (32%) |
| Average | 1,205 (31%) | 290 (30%) |
| High | 1,613 (41%) | 367 (38%) |
| **Alcohol consumption**† |  |  |
| Non-drinker | 1,708 (45%) | 484 (50%) |
| Light drinker | 1,706 (44%) | 370 (38%) |
| Moderate to heavy drinker | 428 (11%) | 113 (12%) |
| **Smoking**‡ |  |  |
| Non-smoker | 2,391 (61%) | 596 (61%) |
| Former smoker | 914 (23%) | 213 (22%) |
| Current, ≤15 cigarettes/day | 355 (9%) | 99 (10%) |
| Current, >15 cigarettes/day | 267 (7%) | 70 (7%) |
| **Leisure time physical activity**¶ |  |  |
| Sedentary | 744 (19%) | 231 (24%) |
| Light | 2,500 (64%) | 615 (63%) |
| Moderate to intense | 650 (17%) | 122 (13%) |
| **Hypertension**§ | 2,632 (67%) | 664 (67%) |
| *Abbreviations: IQR=interquartile range.*  *54 missing values, †131 missing values, ‡35 missing values, ¶78 missing values, §7 missing values | | |

### **Online Supplemental Table 9.** Results of the nested case–control conditional logistic regression analysis

| **Variable** | **OR (95% CI)** | **p-value** |
| --- | --- | --- |
| **History of TBI** |  |  |
| No TBI | 1.0 |  |
| Minor TBI | 0.72 (0.35–1.48) | 0.372 |
| Major TBI | 1.20 (0.74–1.96) | 0.465 |
| **Education status** |  |  |
| Low | 1.0 |  |
| Middle | 0.87 (0.72–1.05) | 0.139 |
| High | 0.84 (0.70–1.01) | 0.070 |
| **Smoking** |  |  |
| Non-smoker | 1.0 |  |
| Former smoker | 0.96 (0.78–1.17) | 0.671 |
| Current, ≤15 cigarettes/day | 1.16 (0.89–1.50) | 0.272 |
| Current, >15 cigarettes/day | 0.98 (0.72–1.34) | 0.912 |
| **Alcohol consumption** |  |  |
| Non-drinker | 1.25 (1.06–1.47) | 0.008 |
| Light drinker | 1.0 |  |
| Moderate to heavy drinker | 1.21 (0.94–1.55) | 0.139 |
| **Leisure time physical activity** |  |  |
| Sedentary | 1.0 |  |
| Light | 0.82 (0.69–0.99) | 0.037 |
| Moderate to intense | 0.69 (0.53–0.89) | 0.004 |
| **Hypertension** | 0.99 (0.85–1.17) | 0.950 |
| *Abbreviations*: CI=confidence interval, OR=odds ratio | | |

### **Online Supplemental Table 10.** Results of the nested case–control conditional logistic regression analysis after excluding controls who later developed dementia

The sttocc function in Stata generates a nested case–control study data by sampling controls from the risk sets. For each case (dementia), the controls (non-dementia) are chosen randomly from participants who are at risk at the failure time of the case. That is, the resulting case–control sample is matched with respect to analysis time—the time scale used to compute risk sets (Langholz, B., and D. C. Thomas. 1990. *Nested case-control and case-cohort methods of sampling from a cohort: A critical comparison*. American Journal of Epidemiology 131: 169–176). Thus, the controls are non-demented at the time of matching but can potentially develop dementia afterwards. Of the 3,952 controls, 433 developed dementia later on. Thus, we conducted another analysis excluding these 433 controls that developed dementia later on, leaving 3,519 controls (non-dementia) and 988 cases (dementia). The results are shown below:

| **Variable** | **OR (95% CI)** | **p-value** |
| --- | --- | --- |
| **History of TBI** |  |  |
| No TBI | 1.0 |  |
| Minor TBI | 0.73 (0.35–1.49) | 0.384 |
| Major TBI | 1.18 (0.71–1.94) | 0.523 |
| **Education status** |  |  |
| Low | 1.0 |  |
| Middle | 0.88 (0.73–1.07) | 0.198 |
| High | 0.82 (0.68–0.99) | 0.039 |
| **Smoking** |  |  |
| Non-smoker | 1.0 |  |
| Former smoker | 0.96 (0.79–1.18) | 0.731 |
| Current, ≤15 cigarettes/day | 1.14 (0.88–1.48) | 0.321 |
| Current, >15 cigarettes/day | 0.95 (0.69–1.30) | 0.741 |
| **Alcohol consumption** |  |  |
| Non-drinker | 1.29 (1.09–1.52) | 0.003 |
| Light drinker | 1.0 |  |
| Moderate to heavy drinker | 1.21 (0.94–1.55) | 0.143 |
| **Leisure time physical activity** |  |  |
| Sedentary | 1.0 |  |
| Light | 0.83 (0.69–0.99) | 0.043 |
| Moderate to intense | 0.69 (0.53–0.89) | 0.005 |
| **Hypertension** | 1.04 (0.89–1.22) | 0.616 |
| *Abbreviations*: CI=confidence interval, OR=odds ratio | | |

### **Online Supplemental Table 11.** Characteristics of participants included in the matched exposed–nonexposed analysis

| **Variables** | **Minor TBI** | | **Major TBI** | |
| --- | --- | --- | --- | --- |
|  | **Exposed**  (n=405) | **Non-exposed**  (n=809) | **Exposed**  (n=280) | **Non-exposed**  (n=584) |
| **Age at baseline**, median (IQR) | 45 (35, 54) | 45 (35, 55) | 53 (44, 60) | 47 (37, 57) |
| **Age at TBI**, median (IQR) | 40 (21, 54) | NA | 54 (40, 67) | NA |
| **Age at dementia**, median (IQR) | 76 (73, 82) | 77 (72, 81) | 72 (66, 76) | 77 (72, 79) |
| **Age at death**, median (IQR) | 71 (59, 78) | 68 (62, 76) | 71 (61, 76) | 72 (64, 79) |
| **Sex** |  |  |  |  |
| Female | 168 (42%) | 341 (42%) | 80 (29%) | 169 (29%) |
| Male | 237 (58%) | 468 (58%) | 200 (71%) | 415 (71%) |
| **Educational status*** |  |  |  |  |
| Low | 137 (34%) | 281 (35%) | 108 (39%) | 222 (38%) |
| Average | 137 (34%) | 270 (34%) | 83 (30%) | 174 (31%) |
| High | 127 (32%) | 252 (31%) | 86 (31%) | 183 (31%) |
| **Alcohol consumption**† |  |  |  |  |
| Non-drinker | 156 (40%) | 318 (40%) | 97 (36%) | 201 (35%) |
| Light drinker | 168 (43%) | 336 (43%) | 113 (41%) | 237 (41%) |
| Moderate to heavy drinker | 69 (17%) | 135 (17%) | 64 (23%) | 136 (24%) |
| **Smoking**‡ |  |  |  |  |
| Non-smoker | 200 (50%) | 404 (50%) | 110 (40%) | 232 (40%) |
| Former smoker | 78 (19%) | 158 (20%) | 54 (19%) | 121 (20%) |
| Current, ≤15 cigarettes/day | 72 (18%) | 139 (17%) | 57 (20%) | 114 (20%) |
| Current, >15 cigarettes/day | 54 (13%) | 106 (13%) | 58 (21%) | 117 (20%) |
| **Leisure time physical activity**¶ |  |  |  |  |
| Sedentary | 100 (25%) | 199 (25%) | 72 (26%) | 145 (25%) |
| Light | 192 (48%) | 387 (48%) | 158 (57%) | 333 (58%) |
| Moderate to intense | 110 (27%) | 219 (27%) | 47 (17%) | 100 (17%) |
| **Hypertension**§ | 181 (45%) | 365 (45%) | 148 (53%) | 317 (54%) |
| *Abbreviations: IQR=interquartile range, TBI=traumatic brain injury*  *18 missing values, †48 missing values, ‡5 missing values, ¶16 missing values | | | | |

### **Online Supplemental Table 12.** Risk of dementia based on the matched exposed–nonexposed Cox proportional hazards analyses

| **Variable** | **HR (95% CI)** | **p-value** |
| --- | --- | --- |
| **History of TBI** |  |  |
| No TBI | 1.0 |  |
| Minor TBI | 0.78 (0.36–1.71) | 0.535 |
| **History of TBI** |  |  |
| No TBI | 1.0 |  |
| Major TBI | 1.66 (0.90–3.07) | 0.106 |
| *Abbreviations*: CI=confidence interval, HR=hazards ratio | | |
